# Proteogenomic analysis of air-pollution-associated lung cancer reveals prevention and therapeutic opportunities

**DOI:** 10.1101/2024.03.11.24304129

**Authors:** Honglei Zhang, Chao Liu, Shuting Wang, Qing Wang, Xu Feng, Huawei Jiang, Yong Zhang, Xiaosan Su, Gaofeng Li

## Abstract

Air pollution significantly impact lung cancer progression, but there is a lack of a comprehensive molecular characterization of clinical samples associated with air pollution. Here, we performed a proteogenomic analysis of lung adenocarcinoma (LUAD) in 169 female never-smokers from the Xuanwei area (XWLC cohort), where coal smoke is the primary contributor to the high lung cancer incidence. Genomic mutation analysis revealed XWLC as a distinct subtype of LUAD separate from cases associated with smoking or endogenous factors. Mutational signature analysis suggested that Benzo[a]pyrene (BaP) is the major risk factor in XWLC. The BaP-induced mutation hotspot, EGFR-G719X, was present in 20% of XWLC which endowed XWLC with elevated MAPK pathway activations and worse outcomes compared to common *EGFR* mutations. Multi-omics clustering of XWLC identified four clinically relevant subtypes. These subgroups exhibited distinct features in biological processes, genetic alterations, metabolism demands, immune landscape, and radiomic features. Finally, *MAD1 and TPRN* were identified as novel potential therapeutic targets in XWLC. Our study provides a valuable resource for researchers and clinicians to explore prevention and treatment strategies for air-pollution-associated lung cancers.

Graphical Abstract

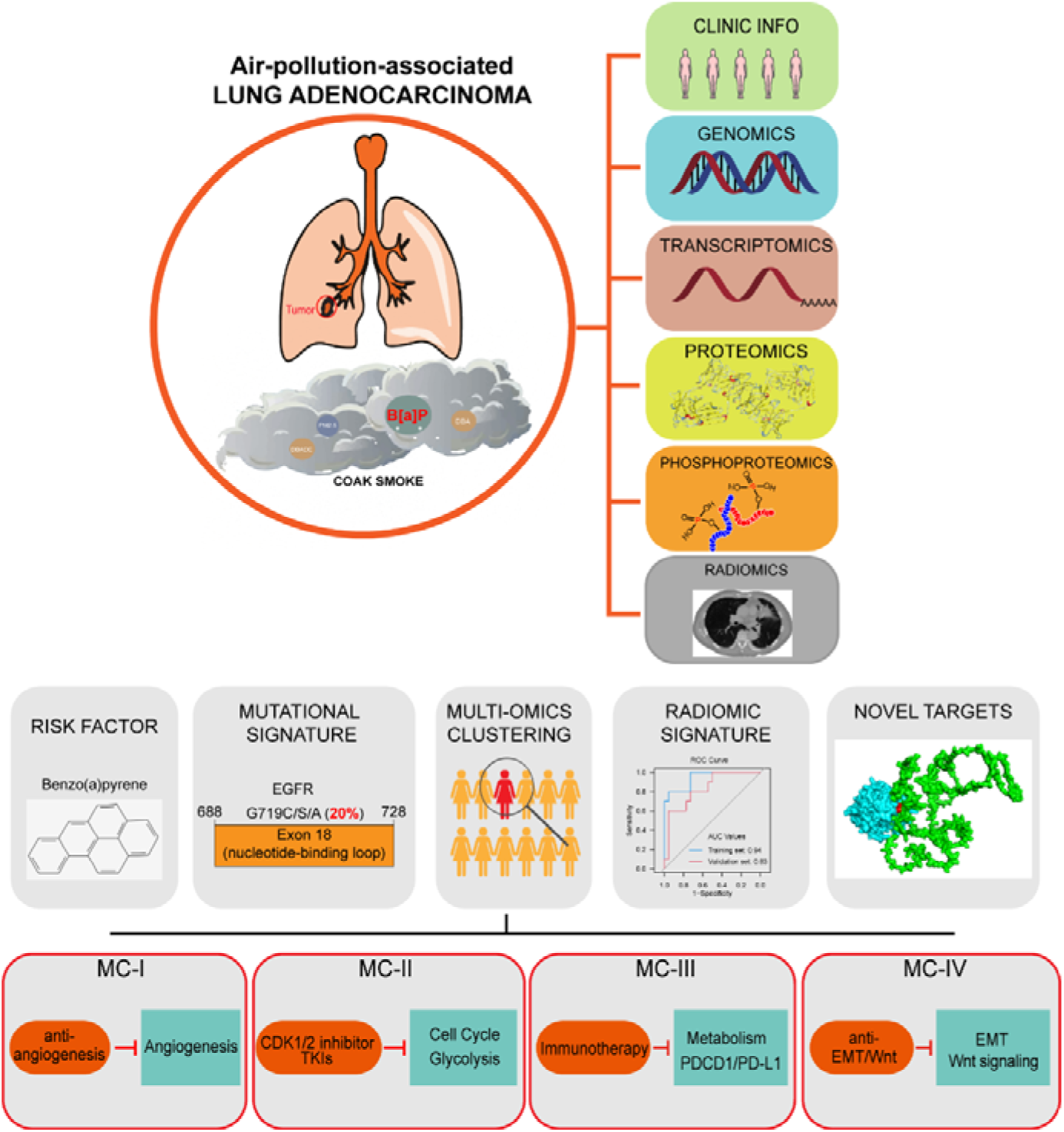

**Highlights:**

- We conducted comprehensive multi-omic profiling of air-pollution-associated LUAD.
- Our study revealed the significant roles of the air pollutant BaP and its induced hot mutation G719X in lung cancer progression.
- Multi-omic clustering enabled the identification of personalized therapeutic strategies.
- Through mutation-informed interface analysis, we identified novel targets for therapeutic intervention.

## Introduction

Lung cancer is the leading cause of cancer deaths globally[1]. Though the most common cause of lung cancer is tobacco smoking, studies estimate that approximately 25% of lung cancers worldwide occur in individuals who have never smoked[2]. Recently, lung cancer in never smokers (LCINS) were molecular profiled and new genomic features were revealed[3–9]. For now, further stratification of LCINS based on different risk factors would be helpful to reveal the oncogenic mechanisms and develop more targeted therapies. Air pollutants, which can directly affect the pulmonary airway, play crucial roles in promoting lung adenocarcinoma[10–12]. More than 20 environmental and occupational agents are lung carcinogens[13, 14] and amount of studies have been made to investigate molecular mechanisms in tumor progression of air pollution chemicals or components using cell lines or mouse/rat models[15–17]. However, a comprehensive molecular characterization of clinical lung cancer samples associated with air pollution is still lacking.

The Xuanwei area has the highest rate of lung cancer in China, and extensive research has established a strong link between lung cancer and exposure to domestic coal smoke[18–23]. Specifically, etiologic link between smoky coal burning and cancer was epidemiologically established[18, 19] and association between household stove improvement and lower risk of lung cancer was observed[23]. Moreover, genomic evidence of lung carcinogenesis associated with coal smoke in Xuanwei area, China was provided in our previous study[22]. Thus, lung cancer in Xuanwei areas exemplifies the ideal disease to study characteristics of lung cancers associated with air pollution. In recent years, the molecular features of Xuanwei lung cancer have been gradually revealed[24–26] [22]. A large sample size with multi-comic molecular profiling is urgent needed to explicit the air pollution chemicals and furthermore propose more targeted therapies.

To better understand the molecular mechanisms and heterogeneity of XWLC and to advance precision medicine, we expanded the sample size of our next-generation sequencing dataset to 169 sample size and performed proteomic and phosphoproteomic profiling to 112 samples. Furthermore, we integrated 107 radiomic features derived from X-ray computed tomography (CT) scans to 115 samples to non-invasively distinguish molecular subtypes. This allowed us to identify potential major risk factor, distinguish the genomic features and establish clinically relevant molecular subtypes. Our study provides an exceptional resource for future biological, diagnostic, and drug discovery efforts in the study of lung cancer related to air pollution.

## Results

To investigate unique biological features of LUAD associated with air pollution, three previous LUAD datasets related to different carcinogens were used for comparison (Fig. 1a). CNLC is the subset of lung adenocarcinoma from non-smoking patients in Chinese Human Proteome Project (CNHPP project) [8] (n=77). TSLC is the subset of lung adenocarcinoma from smoking female in TCGA-LUAD project [27] (n=168). TNLC is the subset of lung adenocarcinoma from non-smoking female in TCGA-LUAD project[27] (n=102). The clinicopathological characteristics of patients from CNLC, TSLC and TNLC cohorts were supplied in Supplementary Table 1a and 1b.

**Fig 1.**
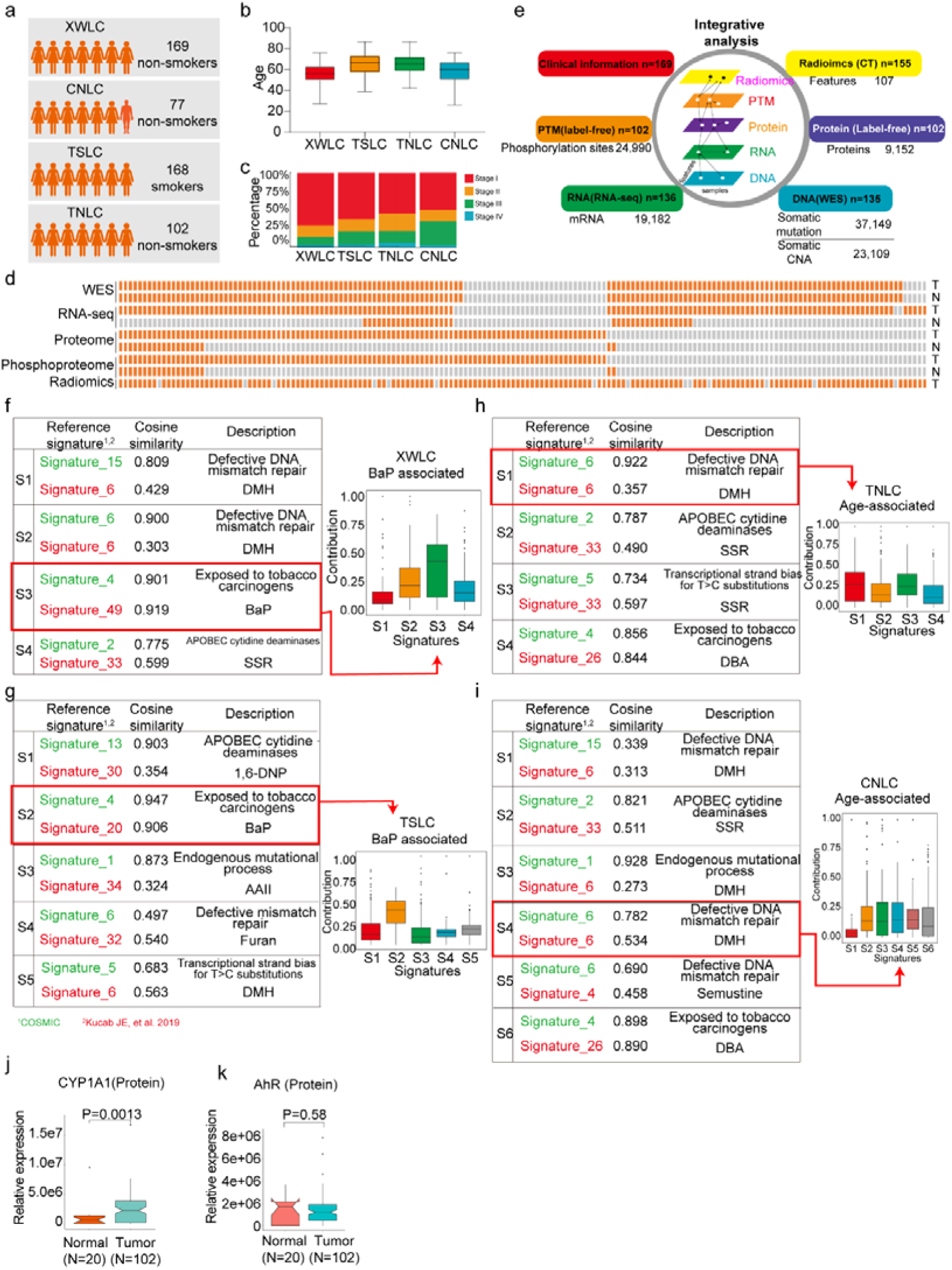
| Proteogenomic profiling and mutational signatures in XWLC. a. Four cohort datasets used in this study: XWLC (Lung adenocarcinoma from non-smoking females in Xuanwei area), CNLC (subset of lung adenocarcinoma from non-smoking patients in Chinese Human Proteome Project), TSLC (subset of lung adenocarcinoma from smoking females in TCGA-LUAD project), TNLC (subset of lung adenocarcinoma from non-smoking females in TCGA-LUAD project); b. Age distribution of patients at the time of operation in the four cohorts; c. Distribution of tumor stages across the cohorts; d. Data availability for the XWLC datasets. Each bar represents a sample, with orange bars indicating data availability and gray bars indicating data unavailability. T, tumor sample. N, Normal tissue; e. Summary of data generated from the XWLC cohort; f-i. Mutational signatures identified in XWLC (f), TSLC (g), TNLC (h), and CNLC (i) cohorts. Cosine similarity analysis of the signatures compared to well-established COMIC signatures (in green) and Kucab et al. signatures (in red). Contribution of signatures in each cohort provided on the right; j-k. Protein abundance of CYP1A1 (j) and AhR (k) in tumor and normal samples within the XWLC cohort; Two-tailed Wilcoxon rank sum test used to calculate p-values in j-k.

## Proteogenomic landscape in Xuanwei lung cancer (XWLC)

The present study prospectively collected primary samples of lung adenocarcinoma (LUAD) from 169 never-smoking women from the Xuanwei area in China (Supplementary Table 1c). The XWLC cohort had a median age of 56 years (Fig. 1b), and the majority of tissue samples were in the early stages of the disease (145 were stage I/II, and 24 were stage III/IV, Fig. 1c). A total of 135, 136, 102 and 102 tumor samples were profiled with whole-exome sequencing (WES), RNA-seq, label-free protein quantification, and label-free phosphorylation quantification, respectively (Fig.1d-e and Extended Fig.1a and 1b). Analysis of the WES data from the paired tumor and normal tissue samples revealed 37,149 somatic mutations, including 1,797 InDels, 32,972 missense mutations, 2,345 nonsense mutations, and 35 nonstop mutations (Supplementary Table 2). Copy number analysis showed 140,396 gene-level amplifications and 67,605 deletions across 40 cytobands (Supplementary Table 3). The mRNA-seq data characterized the transcription profiles of 19,182 genes (Supplementary Table 4). The label-free global proteomics identified 9,152 proteins (encoded by 6,864 genes) with an average of 6,457 proteins per sample (Supplementary Table 5). The label-free phosphoproteomics identified 24,990 highly reliable phosphosites from 5,832 genes with an average of 10,478 phosphosites per sample (Supplementary Table 6). The quality and reproducibility of the mass spectrometry data were maintained throughout the study (Extended Data Fig. 1c-e).

## The air pollutant Benzo[a]pyrene (BaP) primarily contributes to the mutation landscape of XWLC

To infer the primary risk factor responsible for the progression of XWLC, we used SomaticSignatures[28] to identify mutational signatures from single nucleotide variants. Mutational signatures were identified in each cohort and a cosine similarity analysis was performed against mutational signatures in COSMIC mutational signatures[29, 30] and environmental agents mutational sigantures[31] allowing for inference of the underlying causes (Fig. 1f-i and Extended Fig. 2). Generally, exposure to tobacco smoking carcinogens (COSMIC signature 4) and chemicals such as BaP (Kucab signatures 49 and 20) were identified as the most significant contributing factors in both the XWLC and TSLC cohorts (Fig. 1f and 1g). In contrast, defective DNA mismatch repair (COSMIC signature ID: SBS6) was identified as the major contributor in both the TNLC and CNLC cohorts (Fig. 1h and 1i), with no potential chemicals identified based on signature similarities. Therefore, the XWLC and TSLC cohorts appear to be more explicitly associated with environmental carcinogens, while the TNLC and CNLC cohorts may be more associated with defective DNA mismatch repair processes. BaP, a representative compound of polycyclic aromatic hydrocarbons (PAHs), is found in both cigarette smoke and coal smoke and is recognized as a major environmental risk factor for lung cancer[32–34]. Upon metabolism, BaP forms the carcinogenic metabolite 7β,8α-dihydroxy-9α,10α-epoxy-7,8,9,10-tetrahydrobenzo[a]pyrene (BPDE), which creates DNA adducts leading to mutations and malignant transformations. This process involves two key regulators: *CYP1A1* and *AhR*. CYP1A1 plays a crucial role in BaP epoxidation at the 7,8 positions, which is the most critical step in BPDE formation[35]. AhR is a ligand-activated transcription factor that responds to various chemicals, including chemical carcinogens, and is activated by BaP[36]. Accordingly, our results demonstrated significantly higher protein expression of CYP1A1 in tumor samples compared to normal samples (Fig. 1j). AhR showed higher mRNA expression in tumor samples, with no difference in protein level expression (Fig. 1k). All these results suggested the involvement of BaP and its metabolite in the development of lung cancer.

**Fig 2.**
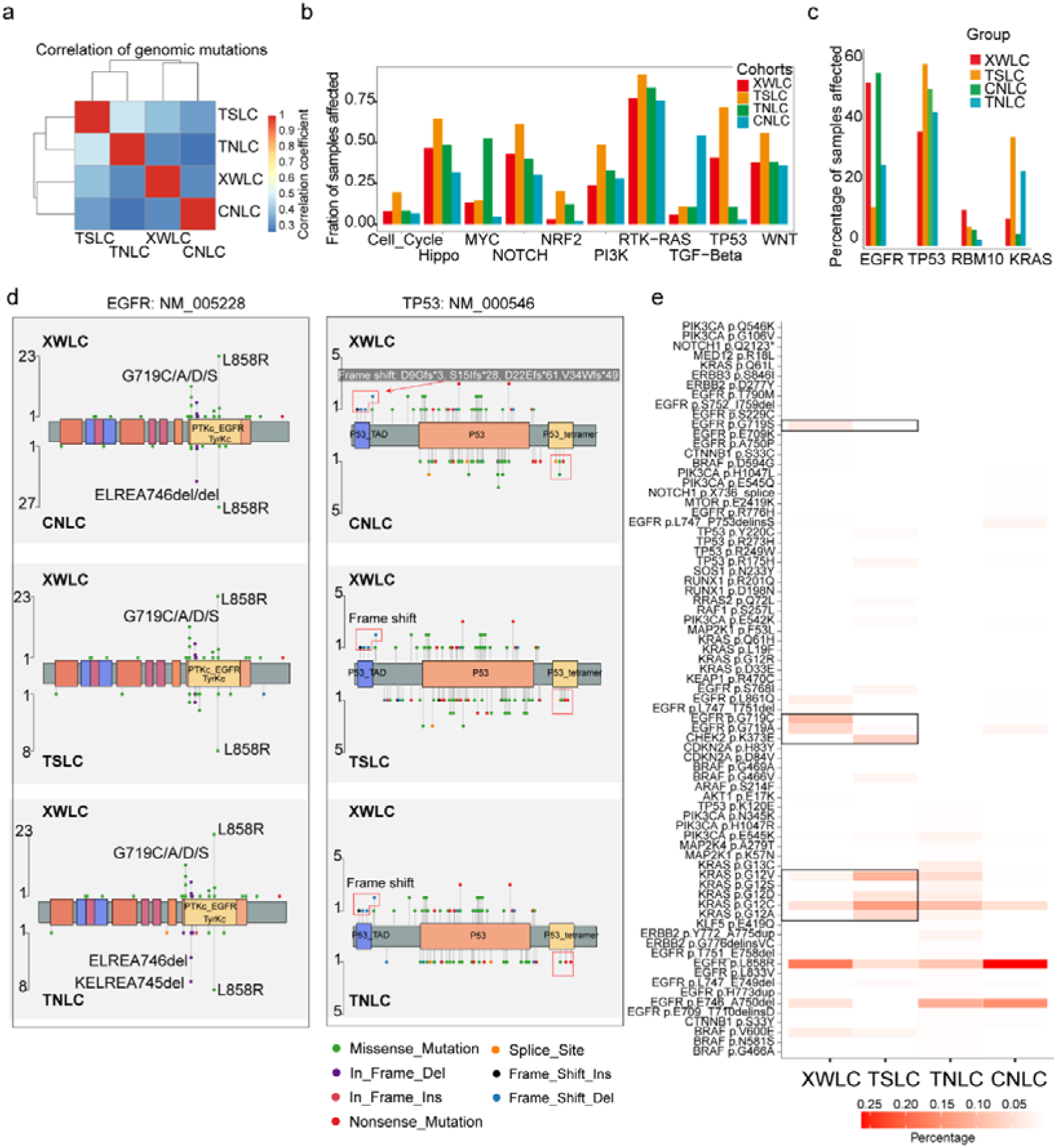
| Genomic and genetic features in XWLC cohort. a. Correlation of genomic mutations among cohorts, determined using the Pearson correlation coefficient; b. Comparison of oncogenic pathways affected by mutations in each cohort; c. Comparison of mutation frequency of four key genes across cohorts; d. Lollipop plot illustrating differences in mutational sites within EGFR (left) and TP53 (right) across XWLC/CNLC, XWLC/TSLC, and XWLC/TNLC pairs; e. Analysis of the percentage of samples with actionable alterations, with a focus on significant variations between XWLC and TSLC cohorts, highlighted by black boxes.

Though coal-smoke related lung cancer (XWLC cohort) and cigarette-smoke related lung cancer (TSLC cohort) showed similar environmental carcinogens, we found that downstream pathway activation and therapeutic targeted potential showed distinctive features. Firstly, the correlation of genomic mutations between XWLC and TSLC was found to be low (Fig. 2a). Secondly, there was a remarkable difference in the fraction of samples affected by pathway mutations between two cohorts (Fig. 2b). Notably, the TSLC cohort exhibited a higher fraction of samples affected by oncogenic pathways comparing to XWLC cohort. Thirdly, mutation frequencies of top mutated genes[22], such as *EGFR*, *TP53*, *RBM10*, and *KRAS* (Fig. 2c), as well as the distribution of amino acid changes in *EGFR* and *TP53*, showed noticeable differences between the XWLC and other cohorts (Fig. 2d). Specifically, the XWLC cohort exhibited a higher mutation rate in G719C/A/D/S within the *EGFR* gene compared to the other three cohorts. For the *TP53* gene, frame shift mutations including D9Gfs*3 (n=1), S15Ifs*28 (n=1), D22Efs*61 (n=1), and V34Wfs*49 (n=2) were exclusively detected in the XWLC cohort, whereas tetramer domain mutations were only found in the other three cohorts (Fig. 2d). Finally, there was a noticeable disparity in the percentage of samples with actionable targets among the cohorts. (Fig. 2e). Actionable targets in XWLC cohort were mainly focus on *EGFR* mutations including pG719S, pG719C, pG719A, pL858R, whereas TSLC cohort had more actionable targets in CHEK2 p.K373E, KRAS p.G12V/D/C/A (Fig. 2e).

Taken together, we found that the XWLC and TSLC cohorts, which are smoke-related lung adenoma groups, demonstrated distinct etiology compared to the TNLC and CNLC cohorts which may be influenced by endogenous risk factors to a greater extent. Additionally, significant disparities were observed between XWLC and TSLC in terms of downstream pathway activations and specific oncogene loci. Consequently, we conclude that air pollution-associated lung cancer represents a distinct subtype within LUAD.

## The EGFR-G719X mutation, which is a hotspot associated with BaP exposure, possesses distinctive biological features

Notably, the XWLC cohort displayed a distinguishable mutation pattern in specific *EGFR* mutation sites compared to the other cohorts (Fig.3a and Fig.2d). In particular, the G719C/A/D/S (G719X) mutation was the most prevalent *EGFR* mutation in the XWLC cohort (20%), while it was rarely found in the other three cohorts (CNLC: 1.9%; TSLC: 1.9%; TNLC: 0) (Fig. 3b). Notably, we found it was a hot spot associated with BaP exposure (Fig. 3c and 3d). Specifically, **GG**C is the 719 codon, the first G can be converted to T (pG719C, n=13) or A (pG719S, n=5), the second G can be converted to A (pG719D, n=1) or C (pG719C, n=8). Thus, pG719C was the most detected mutation type (Fig. 3c). G>T/C>A transversion can be induced by several compounds such as BaP or dibenz(a,h)anthracene (DBA), unlike other compounds, the tallest peak induced by BaP occurs at GpGpG, reflecting how their DNA adducts are formed principally at N^2^-guanine[31]. Our result showed that the most frequently detected pG719C AAchange was correspond to G**G**GC>G**T**GC transversion (Fig. 3d). Thus, pG719C is a hot spot associated with BaP exposure.

**Fig 3.**
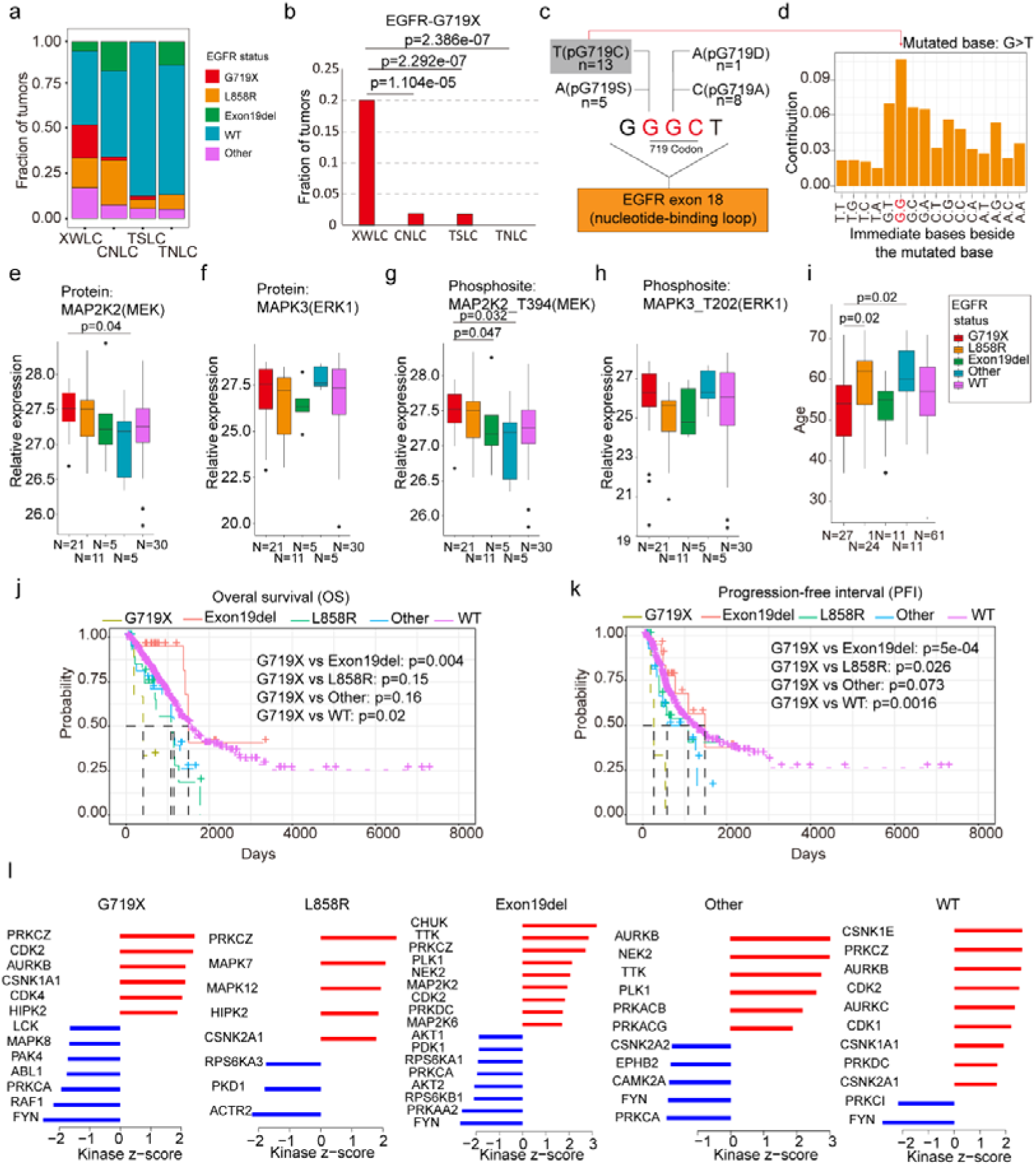
| EGFR-G719X in the XWLC cohort. a. Distribution of different EGFR mutation statuses across the four cohorts; b. Comparison of the fraction of G719X mutations across the four cohorts. Two-sided Fisher’s test was used to calculate *p* values; c. Detailed information on pG719X (pG719/A/D/C/S) mutations in the XWLC cohort. The number of each mutation type is labeled; d. Distribution of nucleotide pairs surrounding the most common G>T transversion site in the XWLC cohort. The x-axis represents the immediate bases surrounding the mutated base. For BaP, the tallest G>T peak occurs at GpGpG; e-h. Comparison of activation levels of key components in the MAPK pathway across different EGFR mutation statuses in the XWLC cohor. N, number of tumor samples containing corresponding EGFR mutation; i. Comparison of patient ages across different EGFR mutation statuses in the XWLC cohort, N, number of tumor samples containing corresponding EGFR mutation; j-k. Presentation of overall survival (OS, j) and progression-free interval (PFI, k) analysis across different EGFR mutation statuses in the TCGA-LUAD cohort, Logrank test was used to calculate *p* values; l. Evaluation of kinase activities by KSEA in tumors across different EGFR mutation statuses in the XWLC cohort. The two-tailed Wilcoxon rank sum test was used to calculate p-values in panels e-i.

We conducted further investigations into the biological characteristics of samples carrying the G719X mutations. Notably, we observed a moderate to high expression of MAPK signaling components, MAP2K2 (MEK), and MAPK3 (ERK1), in tumors harboring the EGFR-G719X mutation compared to other EGFR statuses (Fig. 3e-3h). Utilizing hallmark capability analysis and RNA-seq-based estimation of immune cell infiltration, we found that tumors with G719X mutations exhibited similarities to those with L858R mutations (Extended Data Fig. 3a-b). However, patients with G719X mutations were notably younger than those with L858R mutations, indicating a higher occurrence rate of G719X in younger female patients (Fig. 3i). Analysis of overall survival and progression-free interval (PFI) revealed that patients with the G719X mutation had worse outcomes compared to other EGFR mutation subtypes (Fig. 3j and 3k) which was consistent with a previous study[37]. Furthermore, there were no significant differences in mutation burden or the number of neoantigens between tumors with G719X mutations and tumors with other *EGFR* mutation statuses (Extended Data Fig. 3c).

To explore the heterogeneity of signaling pathways activated by different *EGFR* mutation statuses, we conducted Kinase-Substrate Enrichment Analysis (KSEA) [38, 39] based on the XWLC phosphoproteomics dataset. Our analysis of the phosphoproteome across various *EGFR* mutation types revealed distinct activation patterns of kinases. Specifically, the G719X mutation was associated with the activation of PRKCZ, CDK2, AURKB, CSNK1A1, CDK4, and HIPK2. The L858R mutation showed activation of PRKCZ, MAPK7, MAPK12, HIPK2, and CSNK2A1. The Exon19del mutation exhibited activation of CHUK, TTK, PRKCZ, PLK1, NEK2, MAP2K2, CDK2, PRKDC, and MAP2K6. Other EGFR mutations were associated with the activation of AURKB, NEK2, TTK, PLK1, PRKACB, and PRKACG. EGFR-WT mutations showed activation of CSNK1E, PRKCZ, AURKB, CDK2, AURKC, CDK1, CSNK1A1, PRKDC, and CSNK2A1 (Fig. 3l). In Extended Data Fig. 3d, we provide a list of FDA-approved drugs that target the activated kinases in tumors harboring the G719X mutation.

Currently, afatinib is widely regarded as a first-line therapy for patients with the G719X mutation[40–43]. However, reports indicate that 80% of patients with this mutation may develop resistance to afatinib, even in the absence of T790M[44], underscoring the need for a deeper understanding of the downstream pathways associated with the G719X mutation. Therefore, a promising approach to overcome resistance in tumors with this mutation could involve combining afatinib, which targets activated EGFR, with FDA-approved drugs that specifically target the activated kinases associated with G719X. Therefore, we propose a potential approach to overcoming resistance in tumors with this mutation, which could involve combining afatinib, targeting activated EGFR, with FDA-approved drugs that specifically target the activated kinases associated with G719X.

## Clinically relevant Subtyping in XWLC

To uncover the inherent subgroups within air-pollution-associated tumors, we employed unsupervised Consensus Clustering[45] on integrated RNA, protein, and phosphoprotein profiles of XWLC tumor samples. This analysis led to the identification of four distinct intrinsic clusters, denoted as MC-I, II, III, and IV (Fig. 4a, Extended Data Fig. 4a and Methods). Further survival analysis demonstrated that patients belonging to the MC-IV group exhibited the poorest overall survival compared to the other three subgroups, thus indicating the prognostic potential of multi-omic clustering (Fig. 4b). Notably, there were no significant differences in clinical features such as age and stage observed among the four subgroups (Fig. 4a). As CYP1A1 is a key regulator involved in BaP metabolism and has been proven to be highly expressed in tumor samples (Fig. 1j), we next examined the expression of CYP1A1 among the four subgroups to evaluate their associations with air pollution. Our findings revealed that the MC-II subtype exhibited higher expression of CYP1A1 (Fig. 4c). Moreover, the MC-II possessed more G719X mutations (MC-1:0.39, MC-II:0.42, MC-III: 0.20, MC-IV: 0.08). Notably, there was a significant correlation between CYP1A1 and EGFR expression (Fig. 4e), with EGFR being more highly expressed in the MC-II subtype (Fig. 4e). Collectively, these results indicated that MC-II was more associated with air-pollution.

**Fig 4.**
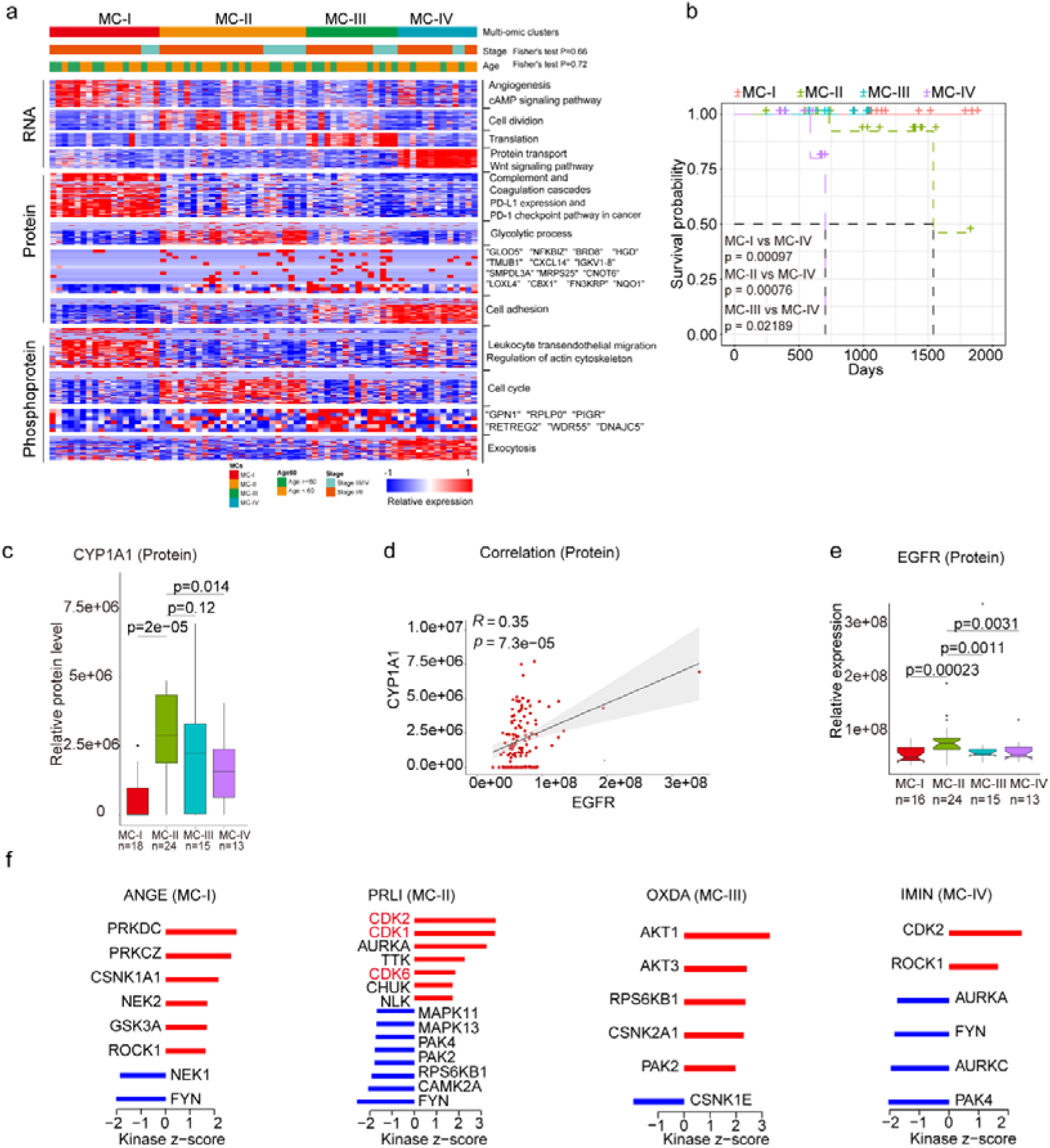
| Subtyping of XWLC. a. Integrative classification of tumor samples into four ConsensusClusterPlus-derived clusters (MC-I to MC-IV). The heatmap displays the top 50 features, including mRNA transcripts, proteins, and phosphoproteins, for each multi-omic cluster. The features are annotated with representative pathways or genes. If a cluster has fewer than 50 features, all features are shown. If no significant GO biological processes are associated with cluster features, all features are displayed; b. Comparison of overall survival between MC-IV and the other three subtypes; c. Protein abundance comparison of CYCP1A1 across subtypes; d. Protein-level correlation between CYCP1A1 and EGFR; e. Protein-level comparison of EGFR across subtypes; f. Evaluation of kinase activities by KSEA in tumors across subtypes in the XWLC cohort. The two-tailed Wilcoxon rank sum test was used to calculate p-values in panels c and e.

Through Kinase Substrate Enrichment Analysis (KSEA) of the phosphoproteome in tumor samples compared to normal adjacent tissues (NATs), we identified specific kinase activations within the four subgroups. In MC-I samples, kinase activations included PRKDC, PRKCZ, CSNK1A1, NEK2, GSK3A, and ROCK1. MC-II samples showed activations of CDK2, CDK1, AURKA, TTK, CDK6, and CHUK. MC-III samples exhibited activations of AKT1, AKT3, RPS6KB1, CSNK2A1, and PAK2. Finally, CDK2 and ROCK1 were activated in MC-IV samples (Fig. 4f). Particularly noteworthy is the enrichment of CDK1/2/6 kinases, which regulate cell cycle checkpoints, in the MC-II subtype, indicating its high proliferation capabilities. These findings imply that distinct kinase pathways are activated within each subgroup, suggesting the presence of specific therapeutic targets for each subgroup. Consequently, we proceeded to explore therapeutic strategies for each subgroup as outlined below:

The **MC-IV** subtype exhibited the poorest overall survival compared to the other three subtypes (Fig. 4b). Given the crucial role of epithelial-mesenchymal transition (EMT) in malignant progression, our first evaluation focused on the EMT process across the four subtypes. We observed higher expression levels of mesenchymal markers such as *VIM*, *FN1*, *TWIST2*, *SNAI2*, *ZEB1*, *ZEB2*, and others in the MC-IV subtype (Fig. 5a). To comprehensively assess the EMT capability of the MC-IV subtype, we calculated EMT scores using the ssGSEA enrichment method based on protein levels and GSEA hallmark gene set (M5930)[46]. The results confirmed the elevated EMT capability of the MC-IV subtype at the protein level (Fig. 5b). Furthermore, Fibronectin (FN1), an EMT marker that promotes the dissociation, migration, and invasion of epithelial cells, was found to be highly expressed in the MC-IV subtype at the protein level (Fig. 5c). Additionally, β-Catenin, a key regulator in initiating EMT, was highly expressed in the MC-IV subtype at the protein level (Fig. 5d). Collectively, our findings demonstrate that the MC-IV subtype is associated with enhanced EMT capability, which may contribute to the high malignancy observed in this subtype.

**Fig 5.**
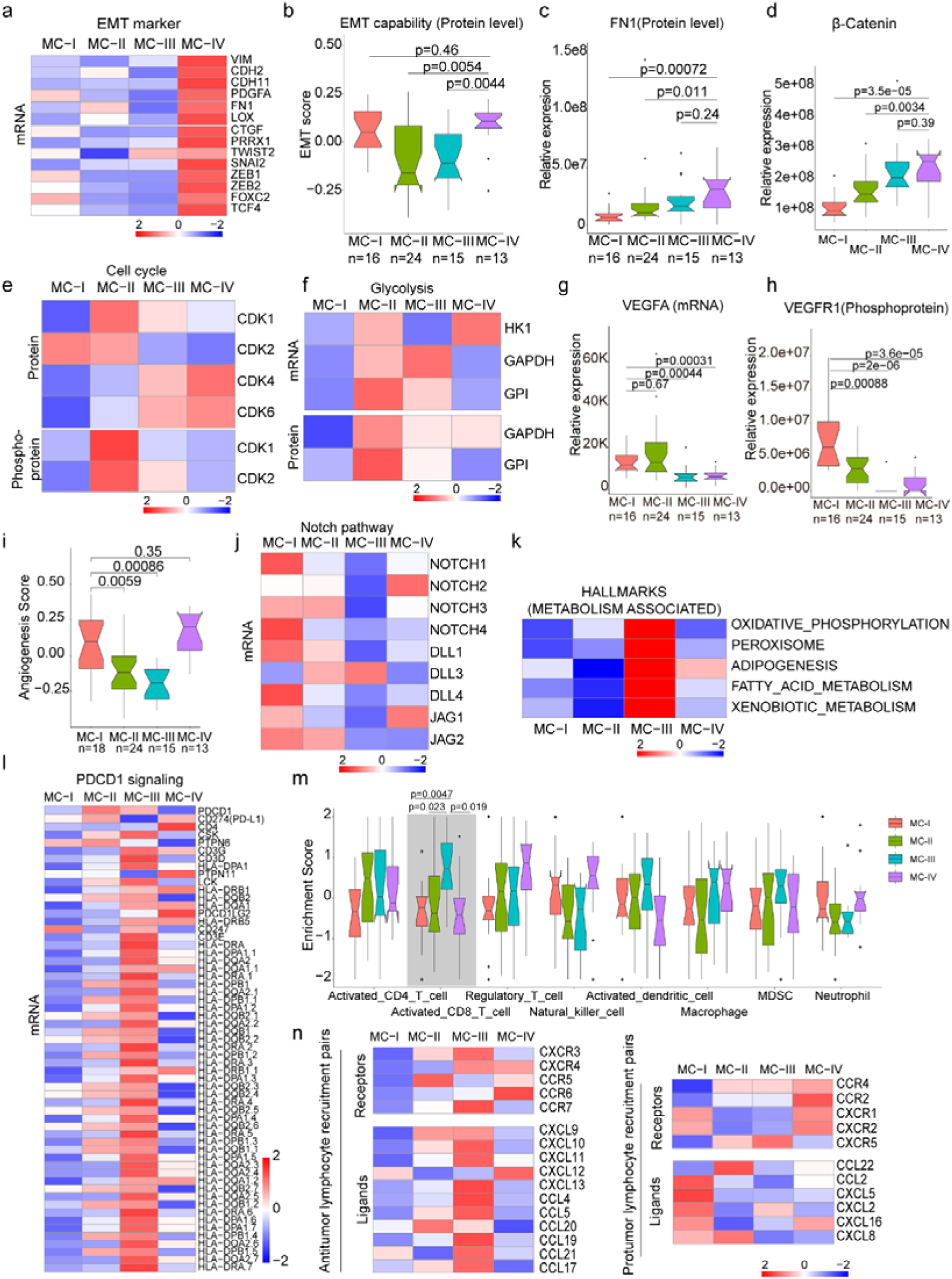
| Biological and immune features across MC subtypes. a. Relative expression of epithelial-mesenchymal transition (EMT) markers across subtypes; b. EMT scores across subtypes using a gene set derived from MsigDB (M5930); c-d. Protein abundance comparison of FN1 (c) and β-Catenin (d) across subtypes; e. Protein and phosphoprotein levels of key cell cycle kinases across subtypes; f. Expression of mRNA and protein levels of glycolysis-associated enzymes; g. mRNA expression of VEGFA across subtypes; h. Phosphoprotein abundance of VEGFR1 across subtypes; i. Angiogenesis score across subtypes using a gene set derived from MsigDB (Systematic name M5944); j. Expression comparison of key regulators of the Notch pathway across subtypes; k. Metabolism-associated hallmarks across subtypes. Gene sets for oxidative phosphorylation, peroxisome, adipogenesis, fatty acid metabolism, and xenobiotic metabolism were derived from MsigDB hallmark gene sets; l. Expression of PD-1 signaling-associated genes across subtypes. PD-1 signaling-associated genes were derived from MsigDB (Systematic name M18810); m. Immune cell infiltration across subtypes. Gene sets for each immune cell type were derived from a previous study[73]; n. Expression of anti-tumor/pro-tumor lymphocyte receptors and ligands across subtypes. The two-tailed Wilcoxon rank sum test was used to calculate p-values in panels b, c, d, g, h, i, and m.

The **MC-II** subtype demonstrated the second-worst outcome and was found to be more strongly associated with air pollution (Fig. 4). This subtype exhibited dysregulation of cell cycle processes, including cell division, glycolysis, and cell cycle biological processes (Fig. 4a). The KSEA analysis revealed that the CDK1 and CDK2 pathways, which are closely linked to cell cycle regulation, were predominantly activated in the MC-II subtype (Fig. 5e). Consistently, we observed higher expression levels of CDK1 and CDK2 at both the protein and phosphoprotein levels in the MC-II subtype, indicating specific elevation of the G2M phase in the cell cycle (Fig. 5e). The cell cycle and glycolysis processes are tightly coordinated, allowing cells to synchronize their metabolic state and energy requirements with cell cycle progression to ensure proper cell growth and division[47–49]. In line with this, we found that key enzymes involved in glycolysis regulation, such as Hexokinase 1 (HK1), Glyceraldehyde-3-phosphate dehydrogenase (GAPDH), and Glyceraldehyde-3-phosphate dehydrogenase-like protein (GPL), were highly expressed in the MC-II subtype (Fig. 5f). Additionally, the MC-II subtype was enriched with EGFR mutations (MC-II vs. others: 18/24 vs. 51/110; Fisher’s exact p = 0.013) and TP53 mutations (MC-II vs. others: 14/24 vs. 35/110; Fisher’s exact p = 0.019), consistent with the characteristic loss of control over cell proliferation. In summary, the MC-II subtype exhibited dysregulated cell cycle processes accompanied by an elevated glycolysis capability, indicating a distinct metabolic and proliferative phenotype.

The **MC-I** subtype exhibited enrichment in various biological processes including angiogenesis, the cAMP signaling pathway, complement and coagulation cascades, PDL1 expression, the PD-1 checkpoint pathway, leukocyte transendothelial migration, and actin cytoskeleton processes (Fig. 4a). In-depth exploration of key components involved in angiogenesis revealed that vascular endothelial growth factor A (VEGFA), a growth factor crucial for both physiological and pathological angiogenesis, was highly expressed in the MC-I subtype (Fig. 5g). Additionally, phosphorylation of vascular endothelial growth factor receptor 1 (VEGFR1), a receptor tyrosine kinase essential for angiogenesis and vasculogenesis, was also highly expressed in the MC-I subtype (Fig. 5h). The angiogenesis scores, calculated using the ssGSEA method based on protein levels and the hallmark gene set (M5944), were relatively high in the MC-I and MC-IV subtypes (Fig. 5i). Furthermore, the relationship between the Notch signaling pathway and angiogenesis is well-established[50]. Notch signaling plays a role in multiple aspects of angiogenesis, including endothelial cell sprouting, vessel branching, and vessel maturation [51, 52]. In the MC-I subtype, the expression of Notch receptors (Notch1-4) and ligands (DLL1, DLL4, JAG1, and JAG2) was highly elevated, indicating increased activation of Notch signaling (Fig. 5j). KEGG pathview analysis demonstrated that key regulators of the VEGF signaling pathway were highly expressed in the MC-I subtype (Extended Fig.5a). Therefore, manipulating Notch signaling could potentially serve as a strategy to regulate angiogenesis and control pathological angiogenesis in the MC-I subtype.

The **MC-III** subtype is characterized by the upregulation of various metabolic processes, including oxidative phosphorylation, peroxisome function, adipogenesis, fatty acid metabolism, and xenobiotic metabolism-related processes (Fig. 5k). Additionally, we conducted further investigations into the immune features across the subtypes. Interestingly, we observed higher expression of genes associated with PD-1 signaling (GSEA, SYSTEMATIC_NAME M18810) in the MC-III subtype (Fig. 5l). Since PD-1 is primarily expressed on the surface of certain immune cells, particularly activated T cells, we inferred the immune cell infiltration using the ssGSEA method based on immune cell-specific gene sets. We found that activated CD8+ T cells exhibited higher infiltration levels in the MC-III subtype compared to the other three subtypes (Fig. 5m and Supplementary Table 7), which may explain the elevated PD-1 signaling in the MC-III subtype. Furthermore, we examined the expression of receptor-ligand pairs involved in both anti-tumor and pro-tumor lymphocyte recruitment. Remarkably, the MC-III subtype exhibited specific high expression of anti-tumor lymphocyte receptors and ligands, while the expression of pro-tumor lymphocyte receptors and ligands was relatively lower (Fig. 5n). In general, the MC-I subtype showed the reverse expression trend in terms of anti-tumor and pro-tumor receptor-ligand pairs (Fig. 5n).

In conclusion, our classification of lung adenocarcinoma associated with air pollution resulted in the identification of four subtypes, each exhibiting distinct biological pathway activation and immune features. The MC-I subtype demonstrated elevated angiogenesis processes, while the MC-II subtype showed a high capacity for cell division and glycolysis. The MC-III subtype exhibited a notable infiltration of CD8+ cells, and the MC-IV subtype was characterized by high EMT capability, which may contribute to its poor outcome. These findings have significant implications for the development of precision treatments for XWLC (Extended Fig.5b).

## Radiomic features across subtypes

Furthermore, we built a noninvasive method to distinguish MC subtypes with radiomics which entails the extensive quantification of tumor phenotypes by utilizing numerous quantitative image features. In the initial step, we defined 107 quantitative image features that describe various characteristics of tumor phenotypes, including tumor image intensity, size, shape, and texture. These features were derived from X-ray computed tomography (CT) scans of 155 patients with XWLC (Methods). The baseline characteristics of this cohort can be found in Supplementary Table 8. Firstly, all features were compared among the four subtypes, and notably, eight features showed significant differences between the MC-II subtype and the other three subtypes (Fig. 6a). Features such as median and mean reflect average gray level intensity and Idmn and Gray Level Non-Uniformity measure the variability of gray-level intensity values in the image, with a higher value indicating greater heterogeneity in intensity values. These results suggest a denser and more heterogeneous image in the MC-II subtype. We further established a signature using a multivariate linear regression model with five image features to distinguish MC-II from the other three subgroups (Extended Fig.6). The performance of the five-feature radiomic signature was validated using the AUC value, which is a generation of the area under the ROC curve. The radiomic signature had an AUC value of 0.94 in the training set and 0.83 in the validation set (Fig. 6b). The confusion matrix revealed an overall accuracy of 0.875 for sample classification using the signature, indicating proficient performance. However, it exhibited suboptimal performance in terms of false-negative classification (Fig. 6c). Taken together, we found that MC-II showed a dense image phenotype, which can be noninvasively distinguished using radiomic features.

**Fig 6.**
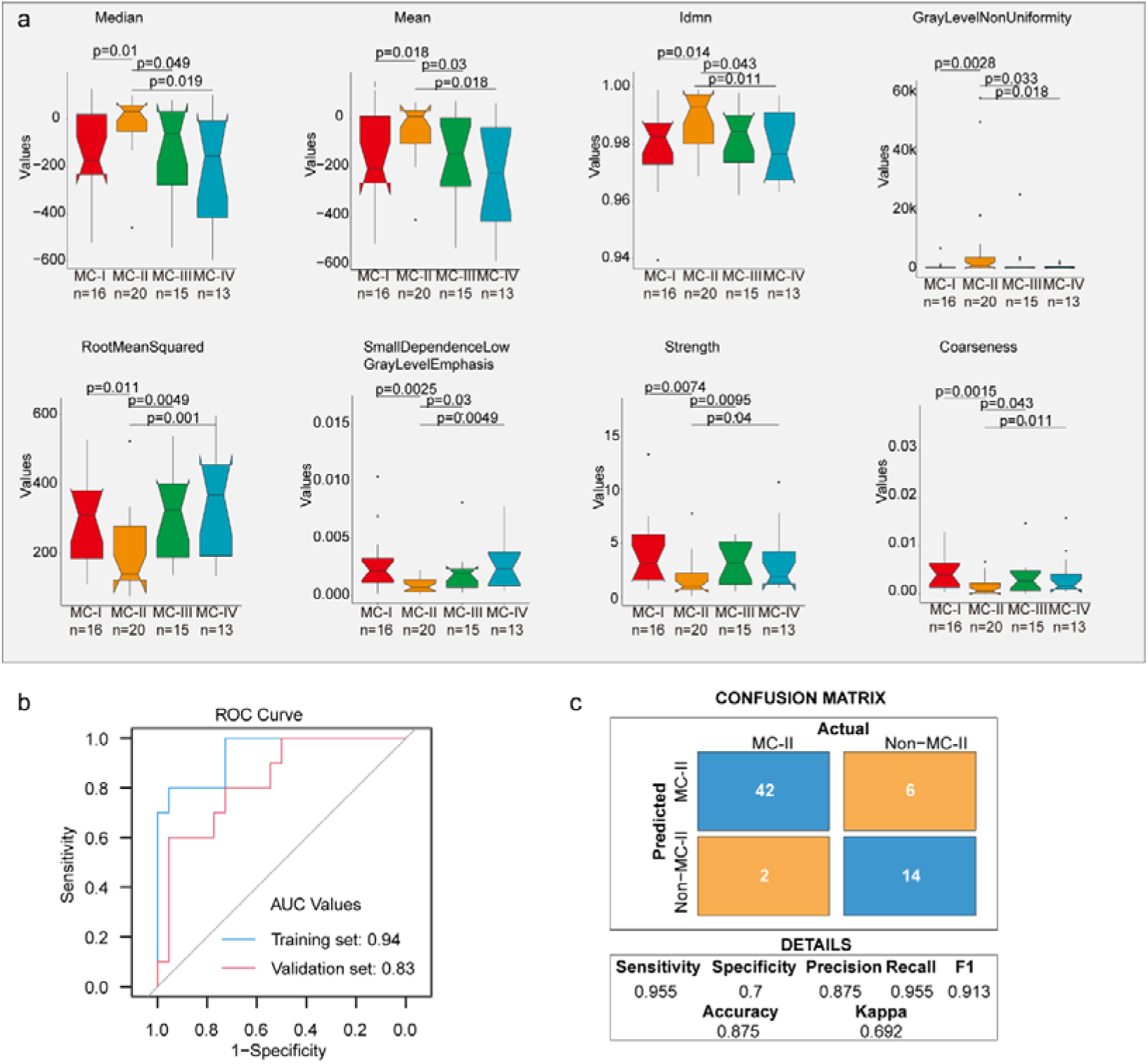
| Radiomic features across subtypes. a. Eight features showing significant differences between MC-II and the other three subtypes. The Wilcoxon rank sum test was used to calculate the p-values; b. A receiver operating characteristic (ROC) curve was used to evaluate the performance of the radiomic signature in distinguishing MC-II from the other three subtypes; c. Confusion matrix allows visualization of the performance of the algorithm in separating MC-II from other subtypes.

## Identification of novel targets based on mutation-informed protein-protein interface (PPI) analysis

The integration of genomics and interactomics has enabled the discovery of functional and biological consequences of disease mutations[53, 54]. To explore novel targets with the concept, we created PPI networks with structural resolution using missense mutations from the XWLC, CNLC, TSLC, and TNLC cohorts (Fig. 7a and Methods). OncoPPIs, defined as a significant enrichment of interface mutations in either of the two protein-binding partners across individuals, were identified in each cohort and were provided in Supplementary Tables 9. The OncoPPIs from the four cohorts are named XWLC_oncoPPIs, CNLC_oncoPPIs, TSLC_oncoPPIs, and TNLC_oncoPPIs, respectively (Fig. 7b and Extended Fig. 7a-c). Initially, the nodes from these four OncoPPIs were subjected to biological process enrichment analysis (Extended Fig. 7d). The analysis revealed that biological processes such as regulation of mitotic cell cycle, TGF-beta signaling pathway, and immune system were predominantly enriched in the genes related to OncoPPIs. Moreover, the processes disrupted by interface mutations showed a relatively higher similarity between the XWLC and TSLC cohorts (Extended Fig. 7d) suggesting convergent targets or pathways affected by smoke-induced mutations.

**Fig. 7.**
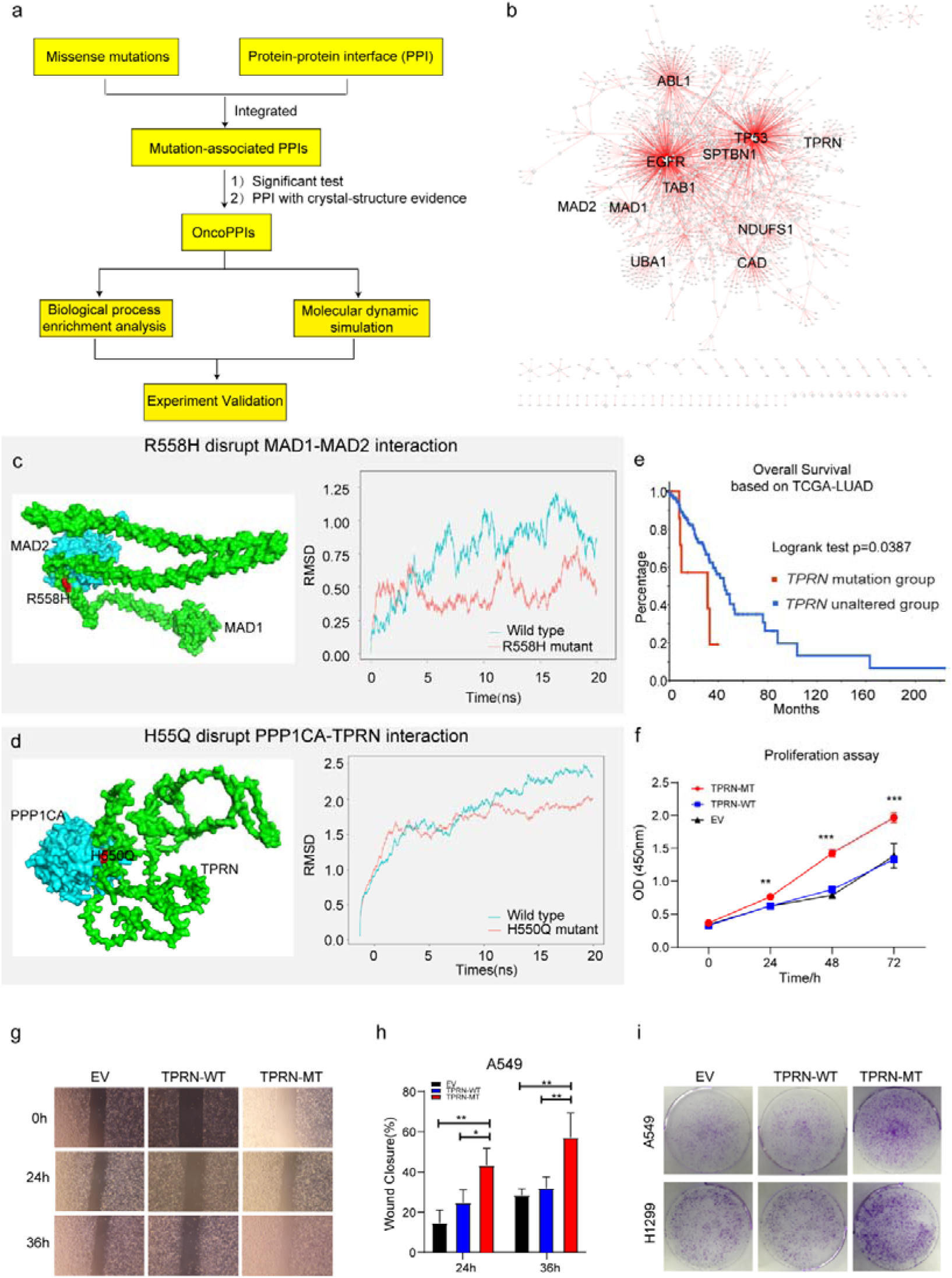
| Identification of novel targets in XWLC. a. Flow chart showing the integration of mutation-informed PPI analysis, molecular dynamic simulation and experiment validation to identify novel targets; b. Network visualization of XWLC_oncoPPIs. Edge thickness represents the number of missense mutations at the protein-protein interaction (PPI) interface, while node size indicates connectivity; c. MAD1-MAD2 interaction model and the p.Arg558His mutation at the interface (left). The complex model was generated using Zdock protein docking simulation. The right distribution showing root-mean-squared deviation (RMSD) during a 20 ns molecular dynamics simulation of MAD1 wild type vs. MAD1 p.Arg558His in the complex; d. Model showing the p.His550Gln alteration within the TPRN-PPP1CA complex (left). The right distribution showing root-mean-squared deviation (RMSD) during a 20 ns molecular dynamics simulation for TPRN wild type vs. TPRN p.His550Gln (H550Q) in the complex; e. Survival analysis of TPRN mutation group and unaltered group derived from cbioportal using TCGA-LUAD cohort (https://www.cbioportal.org/); f. CCK8 assay for empty vector (EV), TPRN-WT and TPRN-MT cell lines in A549 cells which was transfected by EV, TPRN-WT and TPRN-MT, respectively. g. Transwell assay for EV, TPRN-WT and TPRN-MT after 24h and 36h in A549 cells. Magnification was set to 40x; h. Bar chart showing the statistical results of transwell assay; i. Cell colony assay for EV, TPRN-WT and TPRN-MT in A549 and H1299 cell line. The two-tailed Wilcoxon rank sum test was used to calculate p-values in f and h. *, p<0.05; **, p <0.01; ***, p<0.001;

To refine the novel targets from XWLC_oncoPPs, we performed molecular dynamics simulations to predict the binding affinity change by the interface-located mutations (Methods). Mitotic Arrest Deficient 1 Like 1 (MAD1), a crucial component of the mitotic spindle-assembly checkpoint[55, 56], forms a tight core complex with MAD2, facilitating the binding of MAD2 to CDC20[57], which plays a critical role in sister chromatid separation during the metaphase-anaphase transition[58]. Specifically, MAD1 Arg558His has been identified as a susceptibility factor for lung cancer[59] and colorectal cancer[60]. Here, we found that MAD1 allele carrying a p.Arg558His substitution may disrupt the interaction between MAD1 and MAD2 (Fig. 7c). To assess this, we performed molecular dynamics simulations and found that the binding affinity between Arg558His MAD1 and MAD2 was –195.091 kJ/mol and that of wild type was –442.712 kJ/mol (Fig. 7c). Furthermore, on a per-residue basis, the predicted binding affinity (ΔΔG) of Arg558His (Extended Fig. 7e) was projected to increase by 118.319 kJ/mol relative to the wild type (Extended Fig. 7f), indicating that the substitution of Arg558His in MAD1 perturbs the binding affinity. Thus, our findings suggest that the MAD1 Arg558His attributed to lung cancer progression by disrupting of the interaction between MAD1 and MAD2, which showed potential to be explored as a target.

Notably, we identified TPRN as a novel significantly mutated gene in the XWLC cohort (Extended Fig. 7g and 7h) whose status was also associated with patients’ outcomes (Fig. 7e). Previous studies have reported that TPRN interacts with PPP1CA[61]. Thus, we assessed the binding affinity of the TPRN-PPP1CA complex affected by the mutant variant His550Gln. Our result showed that the binding affinities of the complex were –694.372 kJ/mol and –877.570 kJ/mol in mutant and WT cases, respectively (Fig. 7d). On a per-residue basis, the predicted ΔΔG of His550Gln compared to the wild type exhibited an increase of 96.774 kJ/mol (Extended Fig. 7i-j). All these results indicated that TPRN His550Gln increase the binding affinity of the TPRN-PPP1CA complex. To investigate the effect of TPRN His550Gln mutation on tumor progression, we examined proliferation and migration capabilities in both A549 and H1299 lung adenocarcinoma cell lines. CCK-8 assay showed significantly enhanced cell growth after transfection of TPRN mutant allele in both A549 (Fig. 7f) and H1299 cells (Extended Fig. 8a). Moreover, wound-healing assay showed that TPRN mutant cell had achieved enhanced migration capacity (Fig. 7g-h and Extended Fig. 8b-c). Finally, more cell clones in TPRN His550Gln mutation cells were observed in both TPRN-mutant A549 and H1299 cells (Fig. 7i). All these results supported that TPRN His550Gln could be explored as a target in XWLC.

Taken together, our integrated analysis of oncoPPIs and molecular dynamics simulations showed potential to explore novel therapeutic vulnerabilities.

## Discussion

In this study, we conducted proteogenomic and characterized air-pollution-related lung cancers. We found that Benzo[a]pyrene (BaP) influenced the mutation landscape, particularly the EGFR-G719X hotspot found in 20% of cases. This mutation correlated with elevated MAPK pathway activation, worse clinical outcomes, and younger patients. Multi-omics clustering identified four subtypes with unique biological pathways and immune cell patterns. Moreover, our analysis of protein-protein interfaces unveiled novel therapeutic targets. These findings have significant implications for preventing and developing precise treatments for air-pollution-associated lung cancers.

Previously considered uncommon, the EGFR-G719X mutation was detected in only 1-2% of CNLC or TCGA-LUAD cohort samples. Limited knowledge exists from G719X, mostly based on isolated case reports or small series studies[62–66]. In vitro experiments using G719X mutant cell lines and patient-derived xenografts (PDX) demonstrated that osimertinib effectively inhibits signaling pathways and cellular growth, leading to sustained tumor growth inhibition[67]. However, in silico protein structure analysis suggests that G719 alterations may confer osimertinib resistance due to reduced EGFR binding[68]. Presently, afatinib is proposed as the first-line therapy for G719X mutation patients[40–43]. Unfortunately, 80% of G719X patients develop acquired resistance to afatinib without detecting the T790M mutation40. Hence, further mechanistic studies are warranted for G719X. Our study reveals that the G719X mutation is prevalent in the XWLC cohort, significantly impacting treatment selection. Additionally, the large number of G719X samples allowed us to uncover variations in biology and pathway activation, which may facilitate the development of more precise targeted therapies for these patients.

There is substantial evidence linking lung cancer in the Xuanwei area to coal smoke[18, 19, 21, 22, 69, 70]. In addition, we conducted a rat model study that demonstrated the induction of lung cancer by local coal smoke exposure[22]. However, the specific chemical compound in coal smoke responsible for causing lung cancer remains largely unknown. Previous research has mainly focused on studying indoor concentrations of airborne particles and BaP[18, 19, 21, 23, 71]. For instance, studies have shown an association between the concentration of BaP and lung cancer rates across counties[18]. Moreover, improvements in household stoves have led to reduced exposure to benzopyrene and particulate matter, benefiting people’s health[23, 71]. However, these studies primarily relied on epidemiological data, which may be influenced by confounding factors. Mechanistically, Qing Wang showed that BaP induces lung carcinogenesis, characterized by increased inflammatory cytokines, and cell proliferative markers, while decreasing antioxidant levels, and apoptotic protein expression[72]. In our study, we used clinical samples and linked the mutational signatures of XWLC to the chemical compound BaP, which advanced the etiology and mechanism of air-pollution-induced lung cancer. In our study, several limitations must be acknowledged. Firstly, although our multi-omics approach provided a comprehensive analysis of the subtypes and their unique biological pathways, the sample size for each subtype was relatively small. This limitation may affect the robustness of the clustering results and the identified subtype-specific pathways. Larger cohort studies are necessary to confirm these findings and refine the subtype classifications. Secondly, although our study advanced the understanding of air-pollution-induced lung cancer by using clinical samples, the reliance on epidemiological data in previous studies introduces potential confounding factors. Our findings should be interpreted with caution, and further mechanistic studies are warranted to establish causal relationships more definitively. Thirdly, our in silico analysis suggested potential approach to drug resistance in G719X mutations. However, these predictions need to be validated through extensive in vitro and in vivo experiments. The reliance on computational models without experimental confirmation may limit the clinical applicability of these findings.

In summary, our proteogenomic analysis of clinical tumor samples provides insights into air-pollution-associated lung cancers, especially those induced by coal smoke and offers an opportunity to expedite the translation of basic research to more precise diagnosis and treatment in the clinic.

## Declarations

### Ethical Approval

The study protocol was reviewed and approved by the ethical committees of Yunnan Cancer Hospital & The Third Affiliated Hospital of Kunming Medical University (KYCS2022067) and conformed to the ethical standards for medical research involving human subjects, as laid out in the 1964 Declaration of Helsinki and its later amendments.

## Consent to participate

Participants provided written informed consent prior to taking part in the study.

## Consent for publication

Written informed consent was obtained from the patient and the ethical committees of Yunnan Cancer Hospital & The Third Affiliated Hospital of Kunming Medical University for publication of this study.

## Competing interests

The authors declare no competing interests.

## Authors’ contributions

Honglei Zhang, Gaofeng Li, Xiaosan Su and Yong Zhang conceived the study. Honglei Zhang, Chao Liu, Qing Wang and Shuting Wang performed data analysis. Honglei Zhang wrote the manuscript. Xu Feng, Huawei Jiang interpreted the data analysis and drafted the manuscript and critically revised the manuscript. All authors critically revised and gave final approval of the manuscript.

## Funding

This work was supported by the National Natural Science Foundation (Nos. 82273501, 82360613, 82160343, 82060519); Yunnan Basic Research Program (Nos. 202101AZ070001-002, 202401AS070070, 202301AY070001-106); Yunnan Young and Middle-aged Academic and Technical Leaders Reserve Talents Project (No. 202005AC160048); “Famous Doctor” Special Project of Ten Thousand People Plan of Yunnan Province (Nos. CZ0096, YNWR-MY-2020-095); and the Medical Leading Talents Training Program of Yunnan Provincial Health Commission (No. L-2019028).

## Data Availability

Raw sequencing data have been deposited in the Genome Sequence Archive for Human (GSA-Human, https://ngdc.cncb.ac.cn/gsa-human/) under accession codes HRA000124, HRA001481 and HRA001482. Proteomics and phosphoproteomics data have been deposited in the Open Archive for Miscellaneous Data (OMIX, https://ngdc.cncb.ac.cn/omix/) under accession codes OMIX001292. The raw lung CT images used in this paper are available from OMIX under accession codes OMIX002491.

## Acknowledgements

We thank Kesha Yang from Novogene Co., Ltd., Wenfeng Sun from Scale Biomedicine Technology Co., Ltd. (Beijing, China), and Jingli Li from Beijing Qinglian Biotech Co., Ltd. for their help in sequencing and/or bioinformatics analysis. We also thank the “Open and Shared Public Science and Technology Service Platform of Traditional Chinese Medicine Science and Technology Resources in Yunnan” for providing experimental platforms and equipment.

## Notes

### Competing Interest Statement

The authors have declared no competing interest.

### Funding Statement

This work was supported by National Natural Science Foundation (Nos. 82273501, 81960322, 82160343, 82060519, 81860171); Yunnan young and middle-aged academic and technical leaders Reserve Talents Project (No. 202005AC160048); Yunnan basic research program (Nos. 202101AZ070001-002, 202001AY070001-277, 2019FE001(–236)); Medical Reserve Personnel Training Program of Yunnan Provincial Health Commission(No. H-2018097); Famous Doctor Special Project of Ten Thousand People Plan of Yunnan Province(Nos. CZ0096, YNWR-MY-2020-095); Medical Leading Talents Training Program of Yunnan Provincial Health Commission (No. L-2019028).

### Author Declarations

The study protocol was reviewed and approved by the ethical committees of Yunnan Cancer Hospital & The Third Affiliated Hospital of Kunming Medical University (KYCS2022067) and conformed to the ethical standards for medical research involving human subjects, as laid out in the 1964 Declaration of Helsinki and its later amendments.Written informed consent was obtained from the patient and the ethical committees of Yunnan Cancer Hospital & The Third Affiliated Hospital of Kunming Medical University for publication of this study.

### Summary of Updates

Supplemental files updated. Figures revised.

